# Identifying county-level effect modifiers of the association between heat waves and preterm birth using a Bayesian spatial meta regression approach

**DOI:** 10.1101/2025.07.03.25330695

**Authors:** Shuqi Lin, Howard H. Chang, Lyndsey A. Darrow, Matthew J. Strickland, Amy Fitch, Andrew Newman, Xiaping Zheng, Joshua L. Warren

**Affiliations:** Department of Biostatistics, Yale School of Public Health, Yale University, New Haven, CT; Department of Biostatistics and Bioinformatics, Rollins School of Public Health, Emory University, Atlanta, GA; Department of Epidemiology, Biostatistics, and Environmental Health, School of Public Health, University of Nevada, Reno, NV; NSF National Center for Atmospheric Research, Boulder CO, USA

**Author notes:** **Corresponding Author:** Joshua L. Warren, 60 College Street, New Haven, Connecticut 06510.

**Keywords:** conditional autoregressive model, extreme temperature, meta-regression, preterm birth, spatial statistics

## Abstract

High temperature is associated with adverse health outcomes, particularly for vulnerable subpopulations including pregnant individuals and their unborn babies. Several recent studies have investigated the association between temperature and preterm birth at different geographic scales and across different spatial locations. However, there has been less focus on characterizing spatial heterogeneity in risks and identifying modifiable factors that contribute to the observed variation. In this work, we carry out a two-stage modeling approach to (i) estimate county-level short-term associations between heat waves and preterm birth across eight states in the United States and (ii) explore county-level factors that modify these associations using a newly developed hierarchical Bayesian spatial meta-regression approach. Specifically, we extend the traditional meta-regression framework to account for spatial correlation between counties within the same state by modeling the effect estimates using a variant of the conditional autoregressive model. We report several variables that modified the associations between heatwaves and preterm birth, including housing quality, energy affordability, and social vulnerability for minority status and language barriers. An R package, **SpMeta**, is developed for analyses that aims to synthesize area-level risk estimates while accounting for spatial dependence.

## Introduction

High temperatures and extreme heat events are associated with elevated risks of various adverse health outcomes (Anderson et al., 2013; Bell et al., 2024; Bobb et al., 2014). Pregnant individuals and their developing fetuses are potentially more vulnerable to heat exposure due to physiological factors (Bekkar et al., 2020; Ilango et al., 2020). Hence, there is the need to better understand how excessive heat exposure may impact gestational length, such as the risk of preterm birth (i.e., a delivery before 37 completed weeks of gestation). Preterm birth occurs in around 10.5% of all births in the United States (US) and is a major cause of infant mortality and long-term health complications, including respiratory, cognitive, and behavioral outcomes (Bhutta et al., 2002; Ward & Beachy, 2003). Gaining an improved understanding of the role of potentially modifiable environmental factors like temperature is crucial for preventing preterm births and mitigating the impacts of climate change on this vulnerable group.

Current findings, including those from a systematic review and meta-analysis, support that short-term exposures to extreme heat events during pregnancy are associated with an increased risk of preterm birth (Bekkar et al., 2020; Chersich et al., 2020). More recently, Darrow et al. (2024) analyzed more than 53 million births in the 50 largest US metropolitan statistical areas (MSAs) between 1993 and 2017 and found that heat waves were associated with increased rates of preterm birth. A rate ratio of 1.02 (95% confidence interval (CI): 1.00-1.03) was reported for the heatwave metric defined as four consecutive days of temperatures above the local 97.5^th^ percentile, with stronger effects observed among socio-economically disadvantaged groups. Fitch et al. (2025) used a case-crossover analysis of data from eight states across the US during the warm season and similarly found that heat waves were associated with an increased risk of preterm birth, with the strongest associations observed for longer durations of heat wave exposure and higher temperatures relative to local norms.

While Darrow et al. (2024) and Fitch et al. (2025) both used meta-analysis to pool location-specific estimates of the association between heat and preterm birth across MSAs and states, respectively, fewer studies have sought to quantify spatial heterogeneity in risk estimates and assess whether spatially varying factors may explain the observed variability. Meta-regression, an extension of a meta-analysis to include predictors and possibly other random effects, has been widely used in other environmental health settings for this purpose (Bell et al., 2004; Choi et al., 2022; Dominici et al., 2002; Rai et al., 2023). However, the typical statistical frameworks ignore the potential of spatial correlation in the location-specific estimates; a decision which may impact the statistical inference for the included effect modifiers. Exceptions to this have been primarily restricted to the statistical literature, including Sera et al. (2019) who introduced a general mixed-effects meta-regression framework which could accommodate spatial correlation; Wilson et al. (2014) who accounted for point referenced spatial correlation in their second stage meta-regression model of temperature and ozone-related mortality; Sim et al. (2018) who developed a multivariate meta-regression approach using nonparametric Bayesian techniques which included a spatial component to account for residual spatial correlation; and Yu et al. (2021) who extended the existing frameworks to account for functional predictors in addition to spatial correlation.

In this work, we re-analyzed the datasets used in Fitch et al. (2025) at a finer spatial scale with the aim to better characterize spatial risk heterogeneity. Specifically, we first estimate county-level associations between heat wave and preterm birth and then use meta-regression to investigate potential effect modifiers which may contribute to spatial variability. We considered factors related to environmental conditions, socioeconomic status, community resilience, health behaviors, and economic conditions. Additionally, we develop a hierarchical Bayesian meta-regression approach based on disease mapping techniques to address the methodological gap present when conducting meta-regression using areal spatial data. Two versions of the spatial model are developed, applied to our case study, and compared to a traditional non-spatial analysis to better understand the need to account for spatial correlation. Spatially referenced data of this kind are common in environmental epidemiology due to the analysis of large health databases. An R package (i.e., **SpMeta**) is developed so that these models may be applied in future analyses that aims to conduct similar meta-regression of area-level risk estimates.

## Data Description

### Birth and Temperature Data

Birth records from eight US states (i.e., California, Florida, Georgia, Kansas, Nevada, New Jersey, North Carolina, and Oregon) spanning from 1990 to 2017 were collected and used in this analysis, with slight variations due to data availability such that Florida (2004–2017), Georgia (1994–2017), Nevada (1991–2017), and North Carolina (2002–2015) included different years. The analysis was restricted to live, singleton preterm births between 28 and 36 weeks, inclusively. Births were excluded if the ZIP code tabulation area (ZCTA) of the mother was missing, if it could not be linked to a ZCTA from 2010 in the corresponding state, or if the mother delivered outside of the state where she resided.

Temperature data in each state were obtained from the High-resolution Urban Meteorology for Impacts Dataset (HUMID), which aims to better capture urban heat island effects and provides 1 km × 1 km resolution daily maximum and minimum temperature values (Newman et al., 2024). Temperature data were linked to each birth based on the maternal residential address available at the ZIP code level using the 2010 ZCTA shapefile. Heat wave intensity in our study corresponds to the HW3 metric as defined in Darrow et al. (2024). This metric attempts to better reflect heat wave intensity than more standard metrics and is defined as the average degrees (in Celsius) above the 97.5^th^ percentile of the daily average temperature (with the percentile calculated at the ZCTA-level using data from 1990-2017) in the prior four days (including the current days temperature). If the four-day moving average is below the 97.5^th^ percentile, then it is set to zero. Temperatures below the 97.5^th^ percentile are coded as zero during averaging.

### Effect Modifiers

In this study, we utilized a set of county-level predictors to explain county-level variability in the association between heat wave and preterm birth risk, including environmental, socioeconomic, and health factors. Data were obtained from various sources, and Table 1 provides a detailed summary of these predictors, including their descriptions, time periods, and sources.

**Table 1:** Details for the effect modifiers included in the analyses. *BRIC variables were negated so that increasing values indicate worsening conditions with respect to the specific variable.

| Predictor | Label | Description | Time Period | Data Source |
| --- | --- | --- | --- | --- |
| Drought | Drought | Percent of weeks a county is in drought | 1980-present | CDC Environmental Public Health Tracking Network |
| Extreme Precipitation | Precipitation | Annual number of extreme precipitation days | 1990-2017 | NOAA/NCEP North American Land Data Assimilation System |
| Developed Land | Land use | Percent of land used for development | 2001-2016 | National Land Cover Database (NLCD), U.S. Department of the Interior |
| Average Temperature | Temperature | Mean annual temperature aggregated to county level | 1990-2017 | HUMID (High-resolution Urban Meteorology for Impacts Dataset) |
| Summer Maximum Average Temperature | Summer max | Mean maximum temperature during summer months, aggregated to county level | 1990-2017 | HUMID (High-resolution Urban Meteorology for Impacts Dataset) |
| Relative Humidity | Relative humidity | Mean relative humidity aggregated to county level | 1990-2017 | HUMID (High-resolution Urban Meteorology for Impacts Dataset) |
| Obesity Prevalence | Obesity | Prevalence of obesity | 2018 | CDC PLACES Project |
| Physical Activity | Physical activity | Prevalence of no leisure-time physical activity | 2018 | CDC PLACES Project |
| BRIC (Baseline Resilience Indicators for Communities) Index* | Community Economy<br>Environment<br>Housing<br>Institutional<br>Social | Disaster resilience (various categories) | 2010 | Hazards Vulnerability and Resilience Institute, University of South Carolina |
| Energy Affordability | Affordability | Average annual energy burden (% income) | 2018 | The Low-income Energy Affordability Data (LEAD) Tool |
| SVI - Socioeconomic Status | Socioeconomic | Poverty, unemployment, income, and education | 2000 | CDC/ATSDR Social Vulnerability Index |
| SVI - Household Composition & Disability | Household | Age, disability, and single-parent households | 2000 | CDC/ATSDR Social Vulnerability Index |
| SVI - Minority Status & Language | Minority | Race, ethnicity, and English language proficiency | 2000 | CDC/ATSDR Social Vulnerability Index |
| SVI - Housing Type & Transportation | Transportation | Housing structure, crowding, and vehicle access | 2000 | CDC/ATSDR Social Vulnerability Index |
| Percent Below Poverty | Poverty | Percentage of population below poverty line | 2000 | CDC/ATSDR Social Vulnerability Index |

#### Environmental Factors

Several environmental predictors were used to capture the impacts of climate and developed land use. Drought conditions were measured using the percentage of weeks a county experienced drought from 1980 to 2024, as reported by the Centers for Disease Control and Prevention (CDC) Environmental Public Health Tracking Network. Extreme precipitation was measured by the annual average number of extreme precipitation days from 1990 to 2017, as reported by the National Centers for Environmental Prediction (NCEP) North American Land Data Assimilation System. The extent of developed land was measured as the average percentage of land used for development between 2001 and 2016, as reported by the National Land Cover Database (NLCD), which is maintained by the US Department of the Interior (Centers for Disease Control and Prevention, 2024a).

Long-term climate variables, including average temperature, summer maximum average temperature, and summer average relative humidity, were also obtained from HUMID. Mean temperatures on a given day were calculated as the average of the daily maximum and minimum temperatures. Summer maximum temperatures were extracted from the average of the daily maximum temperature for the months of June, July, and August throughout the study period. Summer average relative humidity was estimated using the saturation vapor pressure and daily vapor pressure (computed using dew point temperature) values averaged over June-August throughout the study period. Data were aggregated to the county level by calculating the mean values of grid cells within each county’s boundaries. This method ensures that the climate data reflect average conditions across the entire county rather than at specific point location(Leroux et al., 2000; Newman et al., 2024).

#### Socioeconomic and Resilience Indicators

The Baseline Resilience Indicators for Communities (BRIC) Index, developed by the Hazards Vulnerability and Resilience Institute at the University of South Carolina, provided measures of disaster resilience across six categories (i.e., community, economy, environment, housing, institutional, and social) using data from 2010 (Cutter et al., 2010). We also included the four main themes of the Agency for Toxic Substances and Disease Registry (ATSDR) Social Vulnerability Index (SVI), which include socioeconomic status, household composition and disability, minority status and language, and housing and transportation. Additionally, the percentage of the population below the poverty line was also obtained. All socioeconomic and resilience indicators were measured at the county level, which can offer a comprehensive view of each county’s social and economic conditions (Centers for Disease Control and Prevention; Agency for Toxic Substances and Disease Registry; Geospatial Research, 2000). Energy affordability was measured using the average annual energy burden (as a percentage of income) from the Low-income Energy Affordability Data (LEAD) Tool (Centers for Disease Control and Prevention, 2024a).

#### Health Indicators

Health-related predictors included adult obesity prevalence and physical inactivity rates from 2018, both obtained from the CDC’s Population Level Analysis and Community Estimates (PLACES) Project (Centers for Disease Control and Prevention, 2024b).

## Methods

The first stage of our modeling framework fits county-specific conditional logistic regression models of the preterm birth outcome using individual-level records, applied separately in each county across all eight states included in the study. These county-specific regression analyses are based on a time-stratified case-crossover study design where we only consider the warm season months (May – September), and each case serves as their own controls based on matched control days in the same month during which delivery occurred. Additionally, the model includes a predictor representing the average probability of preterm birth among gestations currently at risk within a person’s ZCTA (Vicedo-Cabrera et al., 2014) and the primary heat wave exposure of interest (i.e., HW3 described in the Birth and Temperature Data subsection) assigned at the ZCTA level. Fitch et al. (2025) used the same average birth probability variable and exposure definition but carried out their analyses at the state instead of county level. Here, we are interested in exploring sub-state variability in these associations and linking the estimated associations with the previously described county-level factors.

Specifically, for county *j* within state *i*, we model the probability of being a case (i.e., a preterm birth outcome) on study day *l* for individual *k* as a function the heat wave metric, adjusting for the average birth probability variable such that

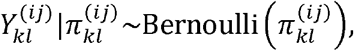

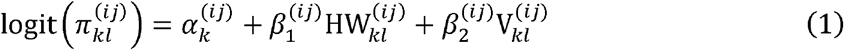

where 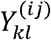 is the binary case/control indicator; 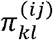 is the probability of being a case; 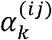 is the intercept specific to individual 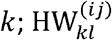 is the HW metric on day *l* with corresponding regression parameter 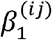; and 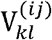 is the average birth probability variable on day *l* in the ZCTA where individual *k* resides with 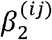 the corresponding regression parameter. In conditional logistic regression, the conditional likelihood function, which excludes the 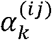 parameters, is used for model fitting in order to reduce bias when the number of strata are large (Avalos et al., 2015). Therefore, these parameters are not actually estimated in our analyses.

In the second stage of our framework, we introduce a hierarchical Bayesian spatial meta-regression model to explore sources of variability in the county-specific HW associations estimated in the first stage while accounting for the clustering of counties within a state as well as correlation between spatially proximal counties in the same state. From the first stage analyses in (1), we obtain estimates of the HW3 association along with the corresponding standard errors (SEs) at each county; 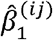 and 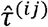, respectively.

Next, as in a typical meta-regression framework, we model these estimates as a function of the true underlying association such that

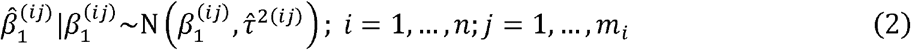

where *n* is the number of states in the study (i.e., 8) and *m*_*i*_ is the number of counties in state *i* (i.e., California, *m*_1_ = 58; Florida, *m*_2_ = 67, Georgia, *m*_3_ = 159, Kansas, *m*_4_ = 101,Nevada, *m*_5_ = 16,New Jersey, *m*_6_ = 21, North Carolina, *m*_7_ = 99, Oregon, *m*_8_ = 34). The true but unobserved association parameter (i.e.,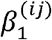) is then modeled as a function of county/state-specific predictor(s) and fixed/random effects to account for clustering and residual spatial correlation such that

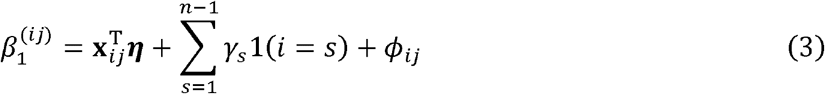

where **x**_*ij*_ is a *p*-length vector of county-specific predictors including an intercept and the variables described in the Data Description section and Table 1, with ***η*** the corresponding set of regression parameters; *γ*_*s*_ is the state *s* intercept parameter with (.) an indicator function and the *n*^th^ state serving as the reference category; and *ϕ*_*ij*_ is the spatially structured random effect term. We consider linear effects as well as nonlinear effects by calculating tertiles for the county-level variables and using those categories as the predictors (with one of the categories serving as the reference group). Additionally, we first fit the model in (2-3) using one predictor at a time before including multiple selected variables in co-adjusted analyses (i.e., those whose 90% credible intervals for *η*_*k*_ exclude zero).

We consider two different approaches for modeling *ϕ*_*ij*_. In both versions, we model the effects from different states independently, as many of our states are geographically separated. In the first version (i.e., Spatial Model 1), we model the collection of random effects from state *i* using the Leroux version of the conditional autoregressive (CAR) model (Leroux et al., 2000) which induces spatial correlation in the random effects based on the neighborhood structure of the counties such that

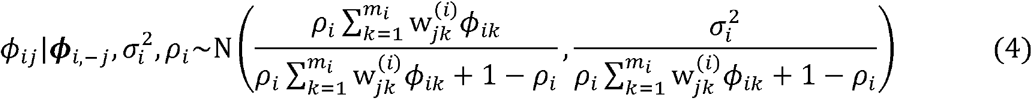

where ***ϕ***_*i,-j*_ is the vector of all *ϕ*_*ik*_ terms connected to state *i* with *ϕ*_*ij*_ removed; 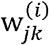 is a binary variable equal to one only if counties *j* and *k* within state *i* share a border, with 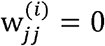 for all *j* by definition; *ρ*_*i*_ ∈ [0,1) describes the strength of spatial correlation between counties in state *i* with zero indicating independence and one representing the original improper intrinsic CAR model (Besag, 1974); and 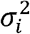 is the state-specific variance parameter.

In the second version of the model for *ϕ*_*ij*_ (i.e., Spatial Model 2), we allow for an additional unstructured error term in order to avoid the possibility of spatial over-smoothing. Specifically, we d efine *ϕ*_*ij*_ = *θ*_*ij*_ + *δ*_*ij*_ where *θ*_*ij*_ are given the same Leroux CAR model detailed in (4) and 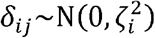 represents the additional non-spatial error term with state-specific variance parameter.

To fully specify the models within the Bayesian setting, we select weakly informative prior distributions for the introduced parameters. Specifically, *η*_1_, …, *η*_*p*_, *γ*_1_,…, *γ*_*n* – 1_ ∼ N(0,100^2^); *ρ*_*i*_∼Uniform(0,1),*i* = 1,…,*n*;and 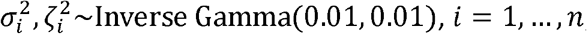; with independence defined between all parameters.

### Model Comparisons

We use Watanabe Akaike information criterion (Watanabe & Opper, 2010), a common Bayesian model comparison tool, to formally compare the different random effect structures across each application of the models. Smaller values of WAIC indicate a model that has an improved balance of fit and complexity. When computing WAIC, we base the likelihood on the marginal distribution of the 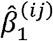 outcomes (i.e., not conditioning on *ϕ*_*ij*_) so that differences in the covariance structures are the primary point of comparison. Specifically, based on (2-3), for Spatial Model 1 we have

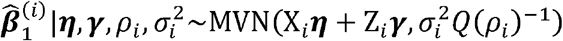

and for Spatial Model 2,

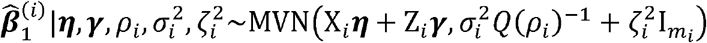

with independence across all states 1,…*n*, where 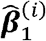 is the vector of 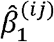 parameters specific to state *i* ; X_*i*_ is an *m*_*i*_ matrix with *J*^th^ row equal to 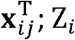 is an *m*_*i*_ by *n* − 1 matrix with *i* ^th^ column equal to all ones for and all other entries equal to zero; Z_*n*_ is a matrix of the same size with all entries equal to zero; *Q*(*ρ*_*i*_) is the precision matrix of the joint distribution of the spatially correlated ***ϕ***_*i*_ parameters whose conditional distribution is defined in (4) (Lee, 2011); and 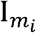 is the *m*_*i*_ by *m*_*i*_ identity matrix. When viewed marginally in this way, Spatial Models 1 and 2 have the same mean and differ only slightly in their covariance structures. For reference, the more traditional non-spatial model has joint distribution defined as 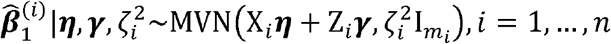. We also fit this model and calculate its WAIC in our analyses to represent the traditional meta-regression technique used widely in the previous literature.

## Results

We fit the county-specific first stage conditional logistic regression models from (1) using proc logistic in SAS 9.4 (SAS Institute Inc., 2013) and estimate the corresponding model parameters. All second stage meta-regression models are fitted in the Bayesian framework using Markov chain Monte Carlo sampling algorithms. An R package to apply these models (i.e., **SpMeta**) is available at https://github.com/warrenjl/SpMeta. Across all models, we let the algorithm run for 110,000 iterations, removing the first 10,000 as burn-in prior to convergence and thinning the remaining 100,000 by a factor of 10 to reduce posterior autocorrelation. Convergence was assessed using visual inspection of individual parameter trace plots and by calculating Geweke’s diagnostic (Geweke, 1991) for each parameter, with no obvious signs of nonconvergence observed across all analyses and parameters. Posterior inference in the form of posterior means and 90/95% quantile-based equal-tailed credible intervals are calculated for exp{*η*_*k*_} parameters. These exponentiated parameters represent the estimated ratio change in odds ratios based on a one standard deviation unit increase in the predictor (i.e., linear analysis) or comparing a selected tertile to the reference tertile (i.e., non-linear analysis). All variables are coded so that increasing values suggest worsening conditions with respect to the specific variable (e.g., increasing poverty, increased social vulnerability).

In Figure 1, we display the raw estimates from the first stage modeling for all counties in the study (i.e.,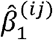), with maps of the SEs (i.e.,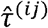) shown in Figure S1 of the Supplemental Material. Some of the counties have missing information because the model in (1) did not converge (8 out of 563, 1.42%). This lack of convergence was due to small sample sizes within a county. Figure 1 shows clear signs of spatial variability in the associations between HW and preterm birth, though a more nuanced interpretation of this figure must factor in the corresponding SEs.

**Figure 1:**
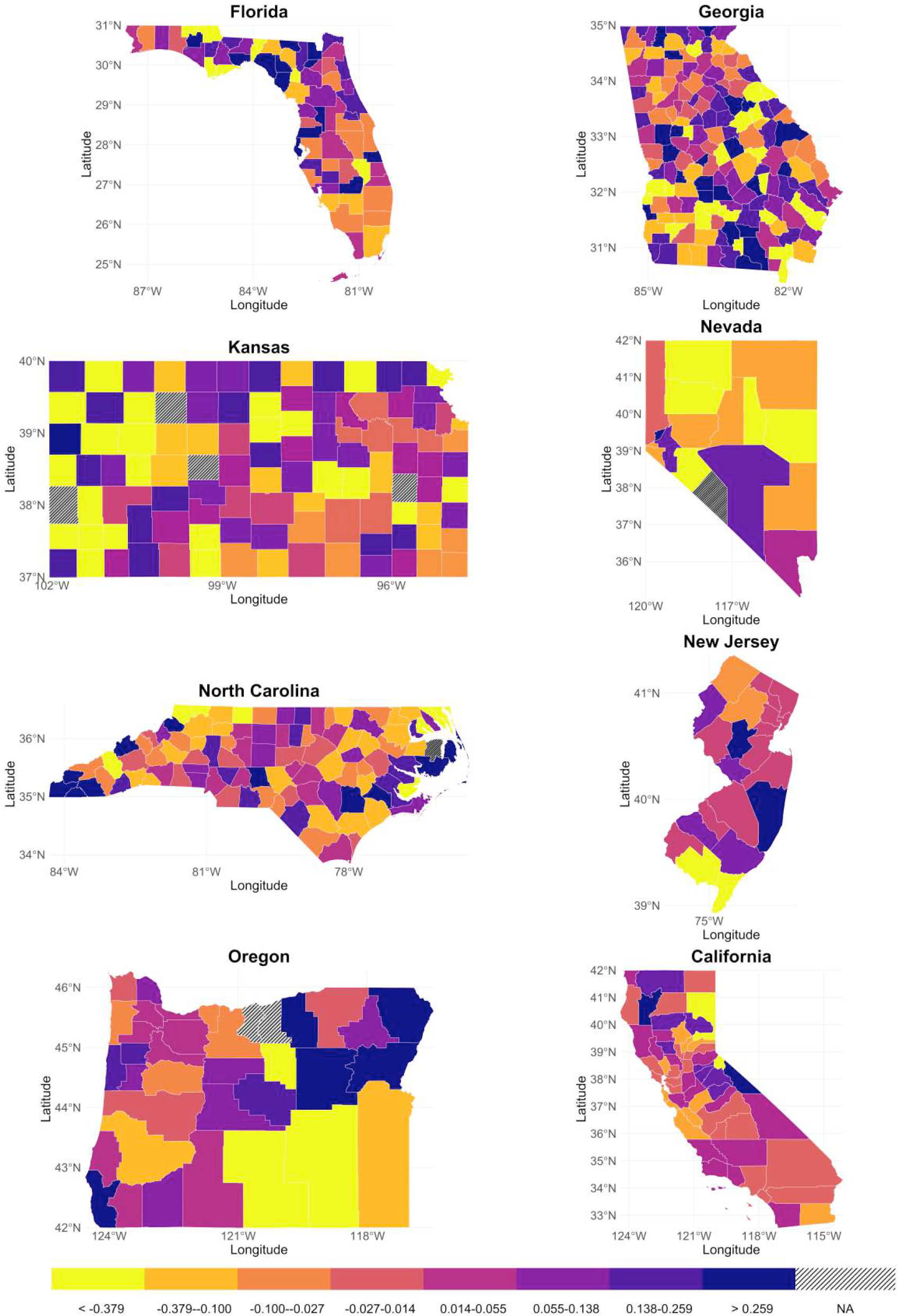
County-level estimates of the association between HW3 and preterm birth from the first stage conditional logistic regression modeling (i.e.,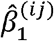) across all states.

In the second stage meta-regression modeling, the WAIC results favored Spatial Model 1 over Spatial Model 2 and the traditional non-spatial model across all of the univariable analyses (see Table S1 of the Supplementary Material). This finding suggests that spatial correlation is present in the first stage estimates and therefore, may be important to account for during analysis. As a result, we present findings from Spatial Model 1 in the main text while displaying results from the other competing models in the Supplemental Material (Tables S2, S3 and Figures S2, S3).

In Table 2 and Figure 2, we display results from the univariable version of Spatial Model 1 for both the linear and non-linear version of the variables. We find evidence that several variables modified the association between heat waves and preterm birth in both the linear and non-linear models. In the linear variable models, higher scores in the economic resilience index (1.012, 90% credible interval (CrI): 1.000–1.024) and housing quality resilience index (1.019, 90% CrI: 1.004–1.034), which suggests worsening conditions for both variables, were associated with a stronger relationship between heat waves and preterm birth (i.e., an increase in the county-specific odds ratios). The social vulnerability index for minority status and language barriers (0.974, 90% CrI: 0.959–0.990) showed an opposite pattern, which suggests areas with higher social vulnerability may have a reduced association. When using 95% Crls, only housing quality resilience index and the social vulnerability index for minority status and language barriers have intervals excluding one.

**Table 2:**
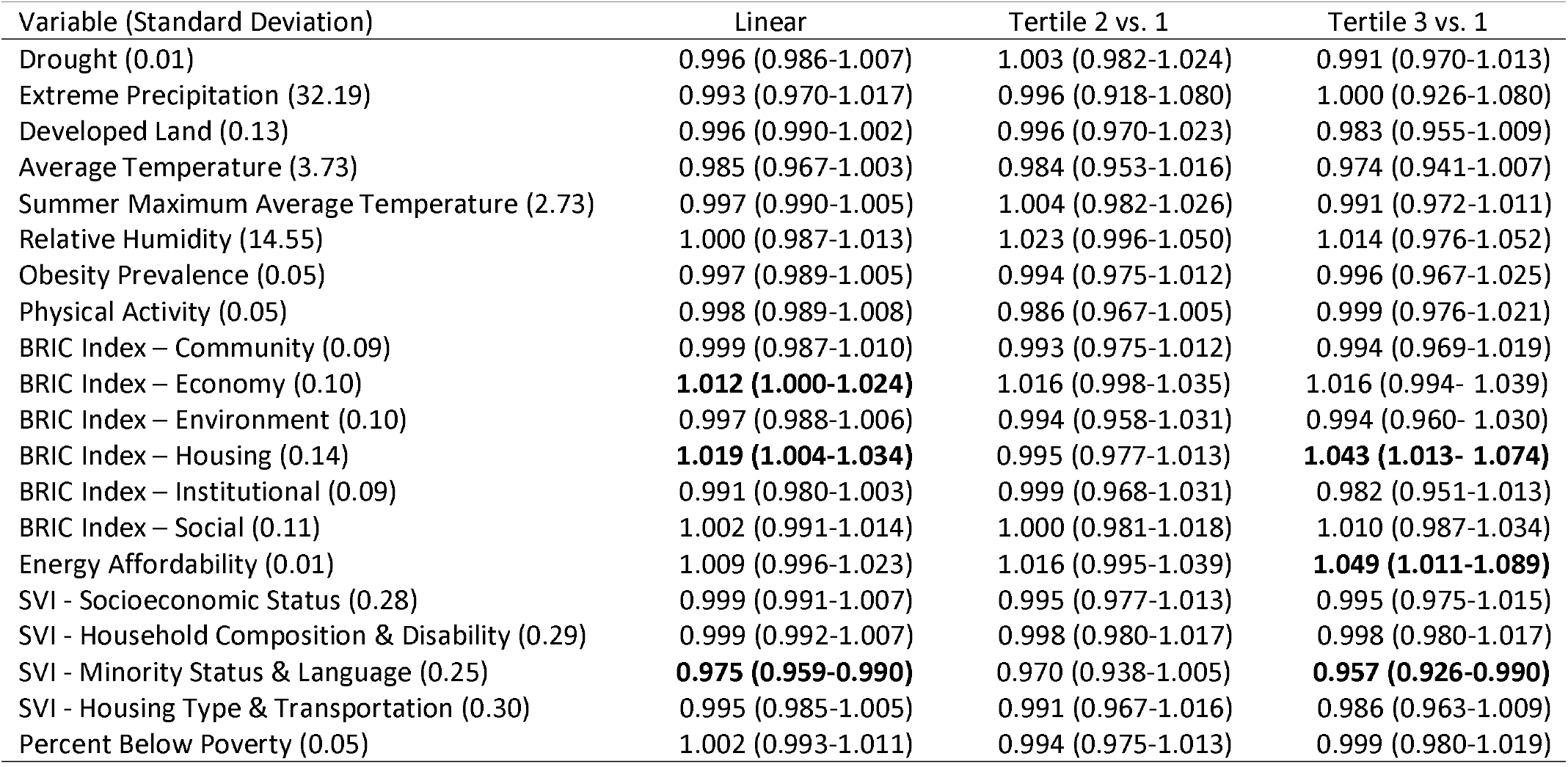
Univariable results based on Spatial Model 1. Posterior means and 90% quantile-based equal-tailed credible intervals are reported for the exponentiated regression parameters from (2) (i.e., exp{*η*_*k*_}) and represent the ratio of odds ratios (from the original analysis between the heatwave metric and preterm birth risk) based on a one standard deviation unit increase in the predictor (i.e., linear analysis) or comparing a selected tertile to the reference tertile (i.e., non-linear analysis). Each variable is coded so that an increasing value in the score represents worsening conditions with respect to the specific variable. Bold entries indicate credible intervals that exclude one.

| Variable (Standard Deviation) | Linear | Tertile 2 vs. 1 | Tertile 3 vs. 1 |
| --- | --- | --- | --- |
| Drought (0.01) | 0.996 (0.986-1.007) | 1.003 (0.982-1.024) | 0.991 (0.970-1.013) |
| Extreme Precipitation (32.19) | 0.993 (0.970-1.017) | 0.996 (0.918-1.080) | 1.000 (0.926-1.080) |
| Developed Land (0.13) | 0.996 (0.990-1.002) | 0.996 (0.970-1.023) | 0.983 (0.955-1.009) |
| Average Temperature (3.73) | 0.985 (0.967-1.003) | 0.984 (0.953-1.016) | 0.974 (0.941-1.007) |
| Summer Maximum Average Temperature (2.73) | 0.997 (0.990-1.005) | 1.004 (0.982-1.026) | 0.991 (0.972-1.011) |
| Relative Humidity (14.55) | 1.000 (0.987-1.013) | 1.023 (0.996-1.050) | 1.014 (0.976-1.052) |
| Obesity Prevalence (0.05) | 0.997 (0.989-1.005) | 0.994 (0.975-1.012) | 0.996 (0.967-1.025) |
| Physical Activity (0.05) | 0.998 (0.989-1.008) | 0.986 (0.967-1.005) | 0.999 (0.976-1.021) |
| BRIC Index – Community (0.09) | 0.999 (0.987-1.010) | 0.993 (0.975-1.012) | 0.994 (0.969-1.019) |
| BRIC Index – Economy (0.10) | **1.012 (1.000-1.024)** | 1.016 (0.998-1.035) | 1.016 (0.994- 1.039) |
| BRIC Index – Environment (0.10) | 0.997 (0.988-1.006) | 0.994 (0.958-1.031) | 0.994 (0.960- 1.030) |
| BRIC Index – Housing (0.14) | **1.019 (1.004-1.034)** | 0.995 (0.977-1.013) | **1.043 (1.013- 1.074)** |
| BRIC Index – Institutional (0.09) | 0.991 (0.980-1.003) | 0.999 (0.968-1.031) | 0.982 (0.951-1.013) |
| BRIC Index – Social (0.11) | 1.002 (0.991-1.014) | 1.000 (0.981-1.018) | 1.010 (0.987-1.034) |
| Energy Affordability (0.01) | 1.009 (0.996-1.023) | 1.016 (0.995-1.039) | **1.049 (1.011-1.089)** |
| SVI - Socioeconomic Status (0.28) | 0.999 (0.991-1.007) | 0.995 (0.977-1.013) | 0.995 (0.975-1.015) |
| SVI - Household Composition & Disability (0.29) | 0.999 (0.992-1.007) | 0.998 (0.980-1.017) | 0.998 (0.980-1.017) |
| SVI - Minority Status & Language (0.25) | **0.975 (0.959-0.990)** | 0.970 (0.938-1.005) | **0.957 (0.926-0.990)** |
| SVI - Housing Type & Transportation (0.30) | 0.995 (0.985-1.005) | 0.991 (0.967-1.016) | 0.986 (0.963-1.009) |
| Percent Below Poverty (0.05) | 1.002 (0.993-1.011) | 0.994 (0.975-1.013) | 0.999 (0.980-1.019) |

**Figure 2:**
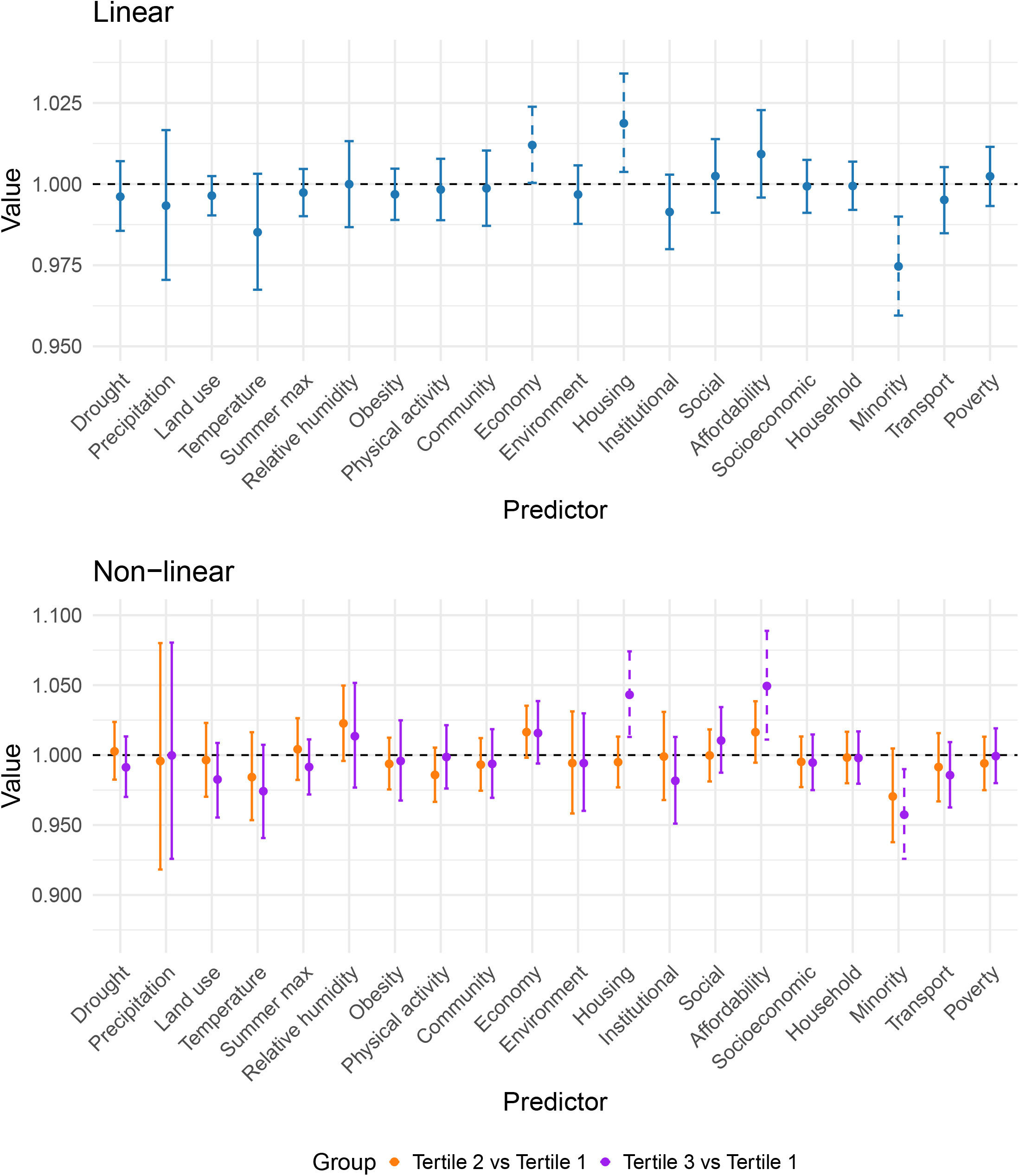
Univariable results based on Spatial Model 1. Posterior means and 90% quantile-based equal-tailed credible intervals are reported for the exponentiated regression parameters from (2) (i.e., exp{*η*_k_}) and represent the ratio of odds ratios (from the original analysis between HW3 and preterm birth risk) based on a one standard deviation unit increase in the predictor (i.e., linear analysis) or comparing a selected tertile to the reference tertile (i.e., non-linear analysis). Each variable is coded so that an increasing value in the score represents worsening conditions with respect to the specific variable. Dashed lines indicate credible intervals that exclude one.

Some variables showed a non-linear pattern in the associations. For the housing resilience index and energy affordability variable, areas in the highest tertile had a stronger association between heat waves and preterm birth compared to the lowest tertile (1.043, 90% CrI: 1.013–1.074 for the housing resilience index; 1.049, 90% CrI: 1.013-1.074 for the energy affordability variable). We observed an opposite trend on the social vulnerability index for minority status and language barriers, where the highest tertile (0.957, 90% CrI: 0.926–0.990) showed a weakening effect on the associations between heatwaves and preterm birth compared to the lowest tertile. When using 95% Crls, housing quality resilience index, social vulnerability index for minority status and language barriers, and energy affordability have intervals excluding one.

Next, we implemented multivariable models by selecting all variables from the univariable results from Spatial Model 1 whose 90% credible excluded one (i.e., energy affordability, social vulnerability index for minority status and language barriers, housing quality resilience index, and economic resilience index). We first checked for correlation among these variables to avoid issues related to multicollinearity and found only low to moderate correlations. We then refit our linear and non-linear versions of the model using Spatial Model 1 while including these significant variables simultaneously (results shown in Table 3). Overall, the results were largely consistent with the univariable findings with effect sizes remaining similar in direction and magnitude.

**Table 3:** Multivariable results based on Spatial Model 1. Posterior means and 90% quantile-based equal-tailed credible intervals are reported for the exponentiated regression parameters from (2) (i.e., exp{*η*_*k*_}) and represent the ratio of odds ratios (from the original analysis between HW3 and preterm birth risk) based on a one standard deviation unit increase in the predictor (i.e., linear analysis) or comparing a selected tertile to the reference tertile (i.e., non-linear analysis). Each variable is coded so that an increasing value in the score represents worsening conditions with respect to the specific variable. Bold entries indicate credible intervals that exclude one.

| Variable (Standard Deviation) | Linear | Tertile 2 vs. 1 | Tertile 3 vs. 1 |
| --- | --- | --- | --- |
| BRIC Index – Economy (0.10) | 1.012 (0.999-1.026) | 1.012 (0.990-1.035) | 0.990 (0.968-1.013) |
| BRIC Index – Housing (0.14) | 1.014 (0.996-1.033) | 1.009 (0.990-1.038) | 0.980 (0.940-1.022) |
| Energy Affordability (0.01) | 0.993 (0.976-1.009) | 1.006 (0.978-1.034) | 1.026 (0.986-1.067) |
| SVI - Minority Status & Language (0.25) | **0.981 (0.964-0.998)** | 1.024 (0.975-1.074) | 0.969 (0.929-1.011) |

## Discussion

In this work, we explored factors that correlate with spatial variability in the association between heat waves and preterm birth by using a two-stage modeling approach. In the first stage we used a case-crossover study design at the county level across eight states to estimate county-specific associations between heat and preterm birth and we found evidence of spatial risk heterogeneity. In the second stage, we introduced a hierarchical Bayesian spatial meta-regression framework to identify effect modifiers while accounting for spatial correlation in the first stage estimates. Using a common Bayesian model comparison tool, we showed that our proposed model performed better in terms of fit and complexity than the traditional non-spatial analysis. The **SpMeta** R package was developed to encourage and facilitate future applications.

Several previous works have investigated the potential for effect modification in the association between preterm birth and extreme heat using interactions and stratified analyses, methods typically applied to a dataset within a single study. Most commonly, these studies have focused on modification by air pollution level, greenspace, and a range of individual-level characteristics, with some evidence of modification often reported (for recent examples, see: (Asta et al., 2019; Cushing et al., 2022; Huang et al., 2021; Ren et al., 2022; Sun et al., 2020; Ye et al., 2024). However, the analyses used in those works differ methodologically from our framework where we are interested in exploring effect modification while pooling findings across multiple analyses/studies (i.e., meta-regression). Less work has been carried out in this setting, with the papers on the topic typically opting for simpler summaries of existing findings or more traditional meta-analysis instead of meta-regression (Bekkar et al., 2020; Darrow et al., 2024; Khosravipour & Golbabaei, 2024; Richards et al., 2022; Sun et al., 2019). An exception is the review from (Chersich et al., 2020), where the authors also carried out a meta-regression though did not account for spatial correlation.

Our findings suggest that counties with lower scores (i.e., improvements) in the economic resilience index, housing quality index, and energy affordability measure, and a higher score (i.e., more vulnerable) in the social vulnerability in minority status and language barriers measure, had a weaker association between heat waves and preterm birth. Most of these findings are intuitive, suggesting increased economic resilience, better housing quality, and improved energy affordability—reflected by lower energy costs relative to household’s total income—mitigate the impact of heatwaves on preterm birth. Improved urban design, architecture, and building design can lessen the impact of extreme heat on individuals as they move within a community setting and during their time spent indoors (Harlan & Ruddell, 2011; Koppe et al., 2004). Interestingly, neighborhoods with a higher proportion of minority populations showed a protective effect against a heat waves’ impact on preterm birth. Similarly, previous studies have reported improved resilience to disasters (e.g., earthquakes, floods) among communities with a higher level of social cohesion (Sobhaninia, 2024). It’s possible that there exists a connection between the social cohesion of a community and its level of minority populations, with respect to extreme heat events, which could at least partially explain this finding. However, further research is needed to fully understand the mechanisms behind this observed protective effect.

Our work has several strengths, including the use of georeferenced birth records data from eight different states across several decades, a consistent framework for generating the first stage estimates of the association between heat waves and preterm birth, a variety of potential effect modifiers from several different data sources, and the development of new statistical methodology and corresponding R package which was shown to outperform the traditional non-spatial meta-regression model. A limitation includes the lack of additional modifiable variables available at the county level (e.g., air conditioning availability). Because we focused on several states that were geographically separated over decades, it was not possible to locate county-level data on every predictor that would have been interesting to include in the framework. However, future work could explore these variables as they become available in different locations and time frames. Additionally, because we focused on areal-level effect modifiers, we were not considering individual-level variables (e.g., maternal age, race). However, the area-level effects could be correlated with some of these potentially important individual-level variables and therefore, the results may reflect effect modification by these factors.

Overall, some of the spatial variability in the association between heat and preterm birth may be explained by certain spatially varying factors. Future work could add to this list of variables, including potentially modifiable features, so that the impact of heat on preterm birth could be mitigated in the future. As the number and intensity of heatwaves continue to increase (U.S. Environmental Protection Agency, 2024), this issue will become increasingly important and mitigation strategies will be needed to protect the most vulnerable subpopulations.

## Data Availability

All data produced are available online at "https://github.com/warrenjl/SpMeta/tree/main/Publication_Data"

https://github.com/warrenjl/SpMeta/tree/main/Publication_Data

