## Supplement for "Identifying county-level effect modifiers of the association between heat waves and preterm birth using a Bayesian spatial meta regression approach"

| Variable | WAIC (pWAIC) | | |
| --- | --- | --- | --- |
|  | Non-Spatial | Spatial Model 1 | Spatial Model 2 |
| Drought | 176.07 (23.60) | **157.81** (21.12) | 224.36 (30.66) |
| Extreme Precipitation | 176.34 (24.03) | **158.70** (21.64) | 225.68 (31.66) |
| Developed Land | 175.76 (24.36) | **158.38** (21.95) | 224.40 (31.67) |
| Average Temperature | 175.20 (23.54) | **156.12** (20.78) | 223.31 (30.85) |
| Summer Maximum Average Temperature | 176.31 (23.57) | **157.84** (20.85) | 224.79 (30.89) |
| Relative Humidity | 176.07 (23.38) | **158.06** (20.74) | 224.60 (30.69) |
| Obesity Prevalence | 175.81 (23.23) | **157.34** (20.58) | 224.22 (30.52) |
| Physical Activity | 176.03 (23.47) | **157.92** (20.81) | 224.46 (30.74) |
| BRIC Index - Community | 176.20 (23.43) | **157.80** (20.72) | 224.46 (30.66) |
| BRIC Index - Economy | 173.46 (23.57) | **155.53** (21.27) | 221.86 (30.82) |
| BRIC Index - Environment | 175.52 (23.53) | **157.52** (20.82) | 224.13 (30.83) |
| BRIC Index - Housing | 171.16 (23.51) | **153.27** (20.64) | 219.06 (30.68) |
| BRIC Index - Institutional | 173.63 (23.32) | **156.06** (20.77) | 222.66 (30.63) |
| BRIC Index - Social | 176.34 (23.89) | **158.63** (21.44) | 224.61 (31.10) |
| Energy Affordability | 174.02 (23.26) | **156.62** (20.76) | 222.33 (30.46) |
| SVI - Socioeconomic Status | 176.11 (23.59) | **158.39** (21.15) | 224.51 (30.84) |
| SVI - Household Composition & Disability | 176.15 (23.62) | **158.33** (21.09) | 224.64 (30.89) |
| SVI - Minority Status & Language | 169.54 (24.46) | **151.07** (21.84) | 218.63 (31.68) |
| SVI - Housing Type & Transportation | 175.17 (23.41) | **157.29** (20.83) | 223.52 (30.71) |
| Percent Below Poverty | 176.00 (23.80) | **158.70** (21.52) | 224.77 (31.18) |
| Table S1: Watanabe Akaike information criterion (WAIC) results for each of the univariable analyses, where smaller values suggest an improved balance of model fit and complexity. pWAIC represents the effective number of parameters in the model, a measure of model complexity. Bold values indicate the best-fitting model. | | | |

| Variable (Standard Deviation) | Linear | Tertile 2 vs. 1 | Tertile 3 vs. 1 |
| --- | --- | --- | --- |
| Drought (0.01) | 1.001 (0.986-1.015) | 1.006 (0.978-1.036) | 0.998 (0.968-1.029) |
| Extreme Precipitation (32.19) | 0.997 (0.970-1.023) | 1.025 (0.929-1.130) | 1.021 (0.928-1.122) |
| Developed Land (0.13) | 0.995 (0.986-1.003) | 0.987 (0.954-1.021) | 0.966 (0.933-1.000) |
| Average Temperature (3.73) | 0.986 (0.963-1.008) | 0.976 (0.938-1.015) | 0.967 (0.925-1.011) |
| Summer Maximum Average Temperature (2.73) | 0.998 (0.988-1.008) | 1.002 (0.971-1.031) | 0.992 (0.963-1.022) |
| Relative Humidity (14.55) | 0.998 (0.980-1.017) | 1.025 (0.989-1.061) | 1.014 (0.969-1.062) |
| Obesity Prevalence (0.05) | 0.999 (0.988-1.010) | 0.993 (0.968-1.018) | 1.003 (0.967-1.040) |
| Physical Activity (0.05) | 1.000 (0.987-1.012) | 0.984 (0.958-1.010) | 0.998 (0.968-1.029) |
| BRIC Index – Community (0.09) | 1.001 (0.987-1.016) | 0.995 (0.969-1.021) | 0.996 (0.965-1.027) |
| BRIC Index – Economy (0.10) | 1.014 (0.999-1.030) | 1.022 (0.995-1.050) | 1.025 (0.994-1.056) |
| BRIC Index – Environment (0.10) | 0.995 (0.984-1.007) | 0.999 (0.957-1.044) | 0.992 (0.952-1.033) |
| BRIC Index – Housing (0.14) | **1.025 (1.007-1.045)** | 0.996 (0.972-1.021) | **1.054 (1.017-1.094)** |
| BRIC Index – Institutional (0.09) | 0.990 (0.975-1.004) | 0.998 (0.962-1.034) | 0.978 (0.942-1.015) |
| BRIC Index – Social (0.11) | 1.004 (0.990-1.018) | 0.995 (0.971-1.020) | 1.014 (0.984-1.046) |
| Energy Affordability (0.01) | 1.013 (0.997-1.029) | 1.019 (0.992-1.047) | **1.051 (1.008-1.094)** |
| SVI - Socioeconomic Status (0.28) | 1.000 (0.989-1.011) | 0.990 (0.966-1.015) | 0.997 (0.970-1.025) |
| SVI - Household Composition & Disability (0.29) | 1.001 (0.991-1.011) | 1.002 (0.976-1.028) | 1.003 (0.977-1.029) |
| SVI - Minority Status & Language (0.25) | **0.973 (0.956-0.991)** | 0.970 (0.931-1.008) | **0.956 (0.919-0.994)** |
| SVI - Housing Type & Transportation (0.30) | 0.993 (0.980-1.006) | 0.988 (0.958-1.019) | 0.982 (0.952-1.012) |
| Percent Below Poverty (0.05) | 1.004 (0.992-1.016) | 0.993 (0.967-1.020) | 1.002 (0.975-1.029) |
| Table S2: Univariable results based on Spatial Model 2. Posterior medians and 90% quantile-based equal-tailed credible intervals are reported for the exponentiated regression parameters from (2) (i.e., $\exp\left\{ \eta_{k} \right\}$) and represent the ratio of odds ratios (from the original analysis between HW3 and preterm birth risk) based on a one standard deviation unit increase in the predictor (i.e., linear analysis) or comparing a selected tertile to the reference tertile (i.e., non-linear analysis). Each variable is coded so that an increasing value in the score represents worsening conditions with respect to the specific variable. Bold entries indicate credible intervals that exclude one. | | | |

| Variable (Standard Deviation) | Linear | Tertile 2 vs. 1 | Tertile 3 vs. 1 |
| --- | --- | --- | --- |
| Drought (0.01) | 0.999 (0.987-1.011) | 1.006 (0.983-1.030) | 0.996 (0.971-1.022) |
| Extreme Precipitation (32.19) | 0.995 (0.976-1.014) | 1.017 (0.932-1.110) | 1.010 (0.929-1.096) |
| Developed Land (0.13) | 0.995 (0.989-1.002) | 0.991 (0.962-1.022) | 0.974 (0.945-1.001) |
| Average Temperature (3.73) | 0.990 (0.972-1.007) | 0.979 (0.947-1.012) | 0.975 (0.940-1.010) |
| Summer Maximum Average Temperature (2.73) | 0.999 (0.991-1.009) | 1.002 (0.977-1.027) | 0.994 (0.971-1.019) |
| Relative Humidity (14.55) | 0.998 (0.984-1.012) | 1.022 (0.993-1.051) | 1.009 (0.971-1.049) |
| Obesity Prevalence (0.05) | 1.000 (0.991-1.009) | 0.997 (0.976-1.018) | 1.003 (0.972-1.034) |
| Physical Activity (0.05) | 1.000 (0.990-1.010) | 0.988 (0.965-1.011) | 0.999 (0.975-1.024) |
| BRIC Index – Community (0.09) | 1.000 (0.990-1.010) | 0.995 (0.974-1.016) | 0.997 (0.973-1.021) |
| BRIC Index – Economy (0.10) | 1.012 (0.999-1.025) | 1.018 (0.996-1.041) | 1.019 (0.994-1.044) |
| BRIC Index – Environment (0.10) | 0.996 (0.987-1.005) | 0.998 (0.963-1.035) | 0.992 (0.959-1.025) |
| BRIC Index – Housing (0.14) | **1.021 (1.005-1.036)** | 0.996 (0.976-1.016) | **1.049 (1.016-1.083)** |
| BRIC Index – Institutional (0.09) | 0.989 (0.977-1.002) | 0.996 (0.964-1.030) | 0.976 (0.944-1.009) |
| BRIC Index – Social (0.11) | 1.002 (0.989-1.014) | 0.996 (0.976-1.018) | 1.010 (0.984-1.036) |
| Energy Affordability (0.01) | 1.011 (0.997-1.024) | 1.017 (0.993-1.040) | **1.047 (1.008-1.087)** |
| SVI - Socioeconomic Status (0.28) | 1.000 (0.992-1.009) | 0.993 (0.973-1.015) | 0.998 (0.975-1.020) |
| SVI - Household Composition & Disability (0.29) | 1.001 (0.993-1.009) | 1.003 (0.982-1.025) | 1.002 (0.980-1.023) |
| SVI - Minority Status & Language (0.25) | **0.975 (0.959-0.991)** | 0.972 (0.937-1.008) | **0.958 (0.924-0.992)** |
| SVI - Housing Type & Transportation (0.30) | 0.994 (0.983-1.006) | 0.990 (0.963-1.017) | 0.984 (0.958-1.010) |
| Percent Below Poverty (0.05) | 1.004 (0.994-1.014) | 0.994 (0.972-1.016) | 1.002 (0.980-1.025) |
| Table S3: Univariable results based on the non-spatial model. Posterior medians and 90% quantile-based equal-tailed credible intervals are reported for the exponentiated regression parameters from (2) (i.e., $\exp\left\{ \eta_{k} \right\}$) and represent the ratio of odds ratios (from the original analysis between HW3 and preterm birth risk) based on a one standard deviation unit increase in the predictor (i.e., linear analysis) or comparing a selected tertile to the reference tertile (i.e., non-linear analysis). Each variable is coded so that an increasing value in the score represents worsening conditions with respect to the specific variable. Bold entries indicate credible intervals that exclude one. | | | |

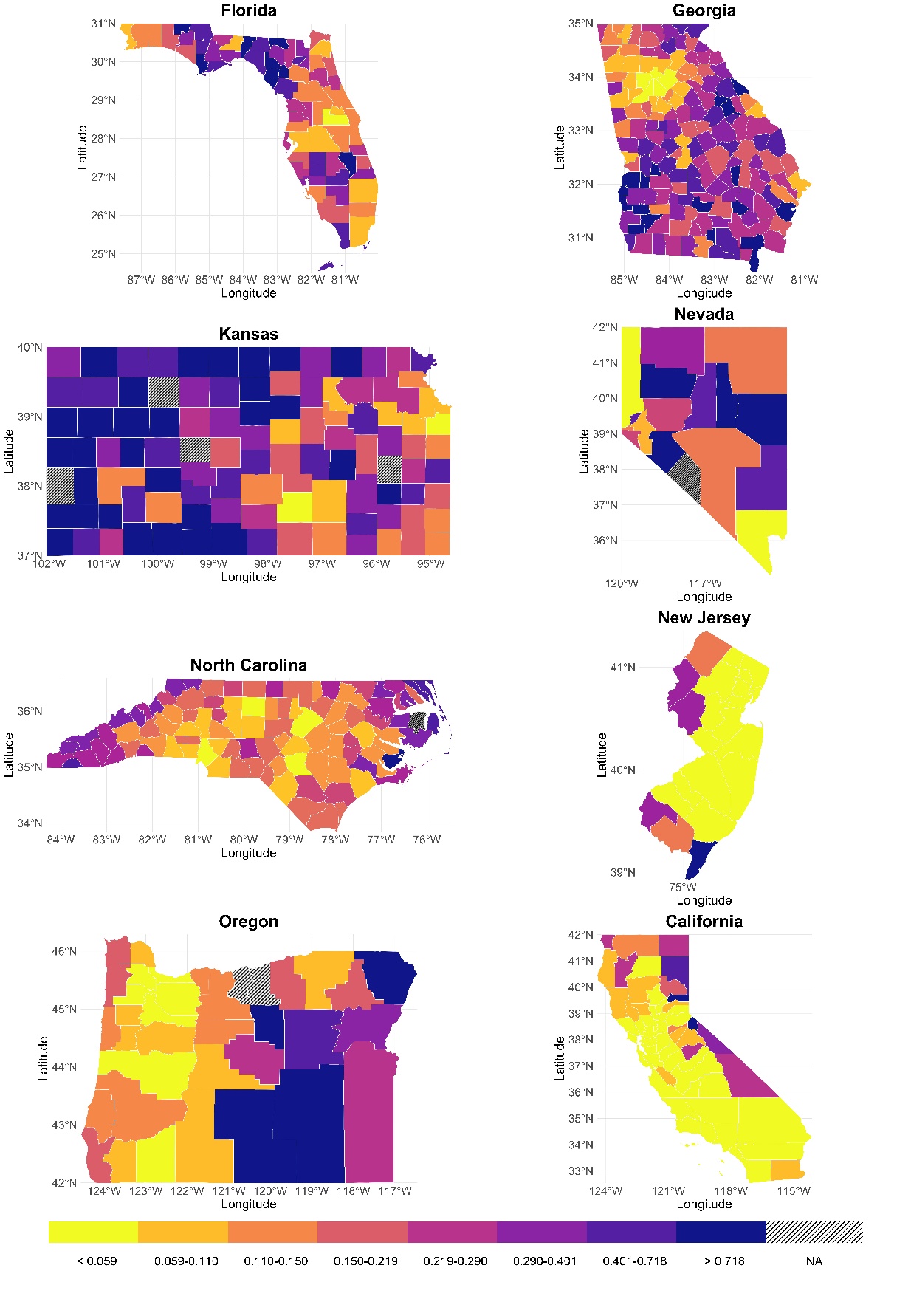

**Figure S1:** County-level standard errors of the estimates of the association between HW3 and preterm birth from the first stage conditional logistic regression modeling (i.e., $\hat{\tau}^{2\left( ij \right)}$) across all states.

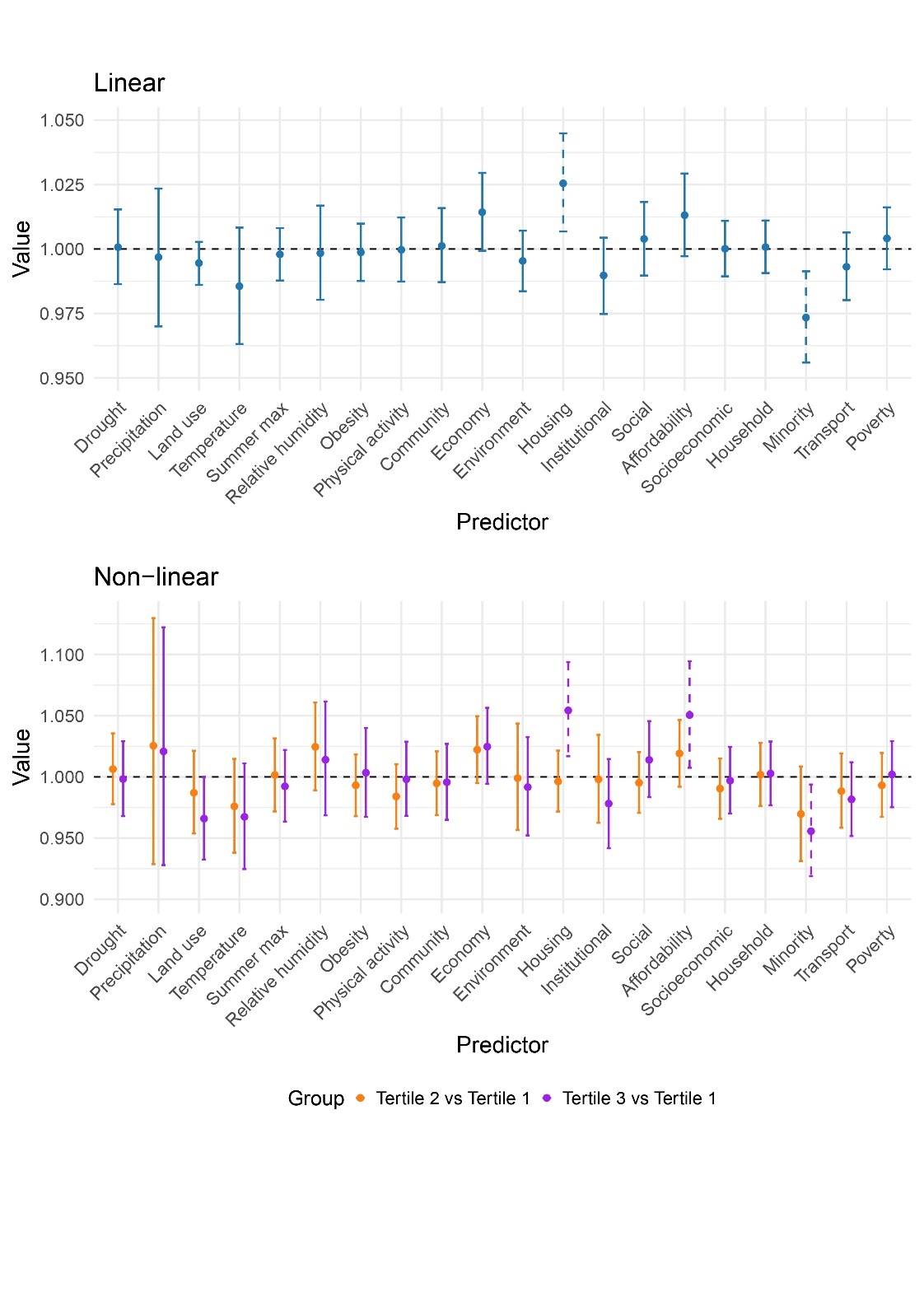

**Figure S2:** Univariable results based on Spatial Model 2. Posterior medians and 90% quantile-based equal-tailed credible intervals are reported for the exponentiated regression parameters from (2) (i.e., $\exp\left\{ \eta_{k} \right\}$) and represent the ratio of odds ratios (from the original analysis between HW3 and preterm birth risk) based on a one standard deviation unit increase in the predictor (i.e., linear analysis) or comparing a selected tertile to the reference tertile (i.e., non-linear analysis). Each variable is coded so that an increasing value in the score represents worsening conditions with respect to the specific variable. Dashed lines indicate credible intervals that exclude one.

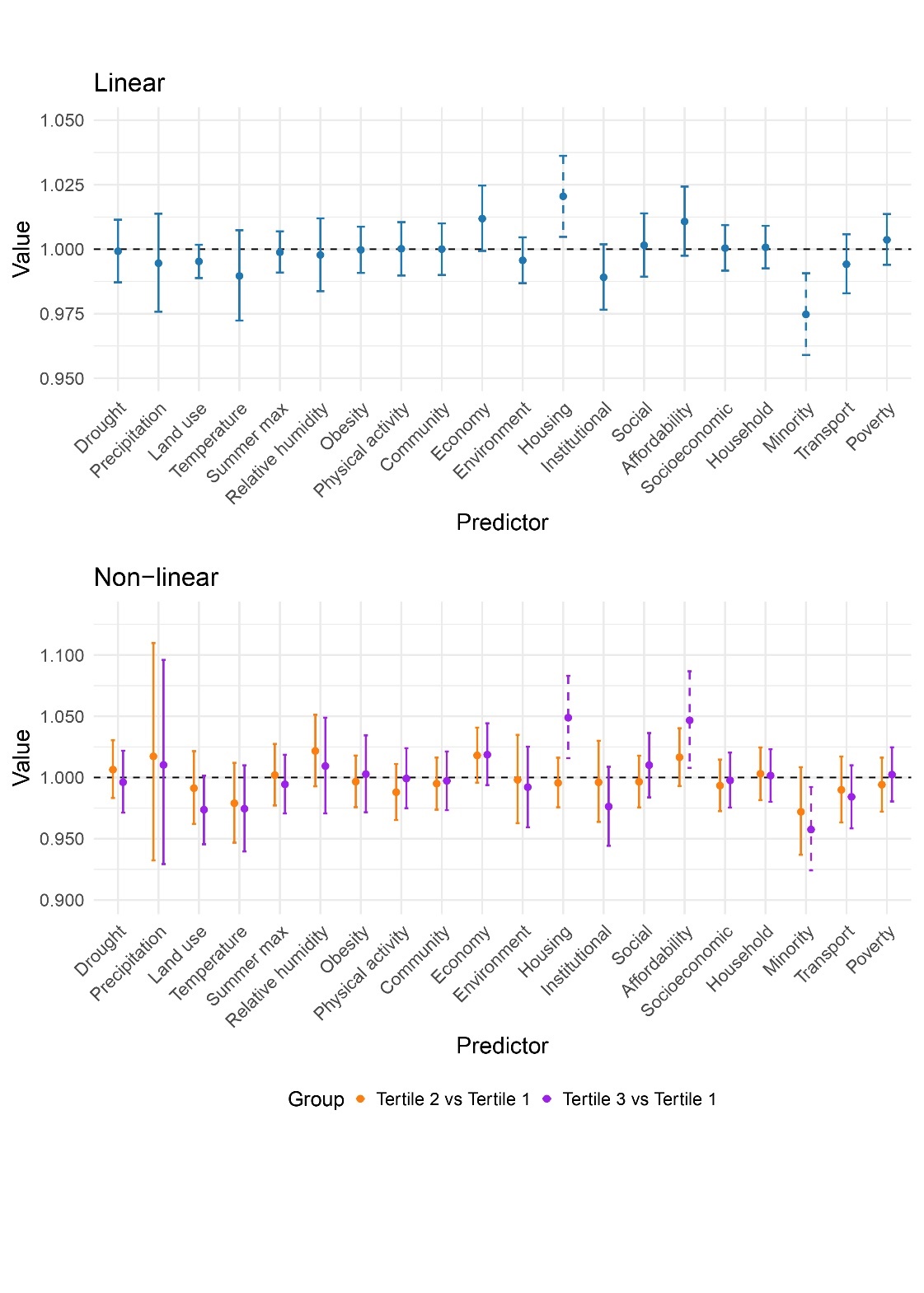

**Figure S3:** Univariable results based on the non-spatial model. Posterior medians and 90% quantile-based equal-tailed credible intervals are reported for the exponentiated regression parameters from (2) (i.e., $\exp\left\{ \eta_{k} \right\}$) and represent the ratio of odds ratios (from the original analysis between HW3 and preterm birth risk) based on a one standard deviation unit increase in the predictor (i.e., linear analysis) or comparing a selected tertile to the reference tertile (i.e., non-linear analysis). Each variable is coded so that an increasing value in the score represents worsening conditions with respect to the specific variable. Dashed lines indicate credible intervals that exclude one.
